# Complement and tissue factor-enriched neutrophil extracellular traps are key drivers in COVID-19 immunothrombosis

**DOI:** 10.1101/2020.06.15.20131029

**Authors:** Panagiotis Skendros, Alexandros Mitsios, Akrivi Chrysanthopoulou, Dimitrios C. Mastellos, Simeon Metallidis, Petros Rafailidis, Maria Ntinopoulou, Eleni Sertaridou, Victoria Tsironidou, Christina Tsigalou, Maria Tektonidou, Theocharis Konstantinidis, Charalampos Papagoras, Ioannis Mitroulis, Georgios Germanidis, John D. Lambris, Konstantinos Ritis

## Abstract

Emerging data indicate that complement and neutrophils are involved in the maladaptive host immune response that fuels hyper-inflammation and thrombotic microangiopathy increasing the mortality rate in coronavirus disease 2019 (COVID-19). Here, we investigated the interaction between complement and the platelet/neutrophil extracellular traps (NETs)/thrombin axis, using COVID-19 clinical samples, cell-based inhibition studies and NETs/human aortic endothelial cell (HAEC) co-cultures. Increased plasma levels of NETs, TF activity and sC5b-9 were detected in patients. Neutrophils yielded high tissue factor (TF) expression and released NETs carrying functionally active TF. Confirming our *ex vivo* findings, treatment of control neutrophils with COVID-19 platelet-rich plasma generated TF-bearing NETs that induced thrombotic activity of HAEC. Thrombin or NETosis inhibition or C5aR1 blockade attenuated platelet-mediated NET-driven thrombogenicity. Serum isolated from COVID-19 patients induces complement activation *in vitro*, which is consistent with high complement activity in clinical samples. Complement inhibition at the level of C3 with compstatin Cp40 disrupted TF expression in neutrophils. In conclusion, we provide a mechanistic basis that reveals the pivotal role of complement and NETs in COVID-19 immmunothrombosis. This study supports emerging strategies against SARS-CoV-2 infection that exploit complement therapeutics or NETosis inhibition.

## Introduction

Accumulated clinical evidence during the evolving pandemic of coronavirus disease 2019 (COVID-19) clearly indicates that severe acute respiratory syndrome coronavirus 2 (SARS-CoV-2) infection can trigger thrombotic complications that affect multiple vital organs, significantly increasing the mortality burden in COVID-19 patients (1–3). The pathological process of coagulopathy in COVID-19 is commonly characterized as an immunothrombosis, since it is related to a maladaptive host immune response that is fueled by excessive activation of innate immune pathways, deregulated inflammation, thrombosis and endothelial dysfunction (2, 4). Understanding the immunothrombotic mechanisms in COVID-19 constitutes a significant medical challenge.

Increased neutrophil count has been associated with disease severity and poor clinical outcomes in COVID-19, and extensive neutrophil infiltration of pulmonary capillaries has been described in autopsy specimens (5–9). In several inflammatory disorders (10), experimental and clinical studies have already demonstrated that neutrophil extracellular traps (NETs) can exert thrombogenic activity through the expression of functionally active tissue factor (TF). NETs appear to be involved in COVID-19 as well (11).

Over-activation of complement has been implicated as an early driver of the maladaptive inflammatory response in COVID-19. Complement can enhance neutrophil/monocyte activation and recruitment to the epithelium of the infected lungs, and several complement effectors, acting in concert with platelets, can fuel thromboinflammation, microvascular thrombosis and endothelial dysfunction (thrombotic microangiopathy) in patients (9, 12). Therefore, early clinical data have prompted the initiation of trials to evaluate various complement-inhibitory strategies in COVID-19 patients (13).

The well-established cross-talk between complement and neutrophils in immunothrombosis in human disorders (14–17) has prompted us to hypothesize that the collaboration of these innate immune systems may mediate early events leading to coagulopathy in COVID-19. Here, we have investigated the role of neutrophils in the thromboinflammation associated with COVID-19 and now provide evidence that complement activation is a key mediator in the activation of the platelet/NETs/TF/thrombin axis during SARS-CoV-2 infection.

## Results and discussion

### NETs are associated with activation of the TF/thrombin axis in COVID-19

We first used ELISA to measure the levels of myeloperoxidase (MPO)/DNA complexes (Figure 1A), a well-defined surrogate marker of NETosis (10), in the plasma of control and COVID-19 patients and detected significantly increased levels in the plasma obtained from COVID-19 patients. The levels of these complexes were positively correlated with thrombin-antithrombin (TAT) activity, suggesting that activation of the TF/thrombin axis had occurred (Figure 1B,C). These findings were further confirmed *ex vivo* by confocal immunofluorescence microscopy in neutrophils collected from four patients with severe COVID-19 (Supplemental Table S1). We found spontaneous formation of NETs expressing TF (Figure 1D,E) that were functional, as indicated by TAT assay (Figure 1F). Moreover, the concomitant increase in TF mRNA levels (Figure 1G) further confirmed previous studies indicating that neutrophils constitute an active source of TF in thromboinflammation. Although detection of NETosis markers in plasma by similar laboratory techniques has already been reported (11), we now show that NET release is positively correlated with *in vivo* thrombotic potency in COVID-19 (Figure 1C).

**Figure 1:**
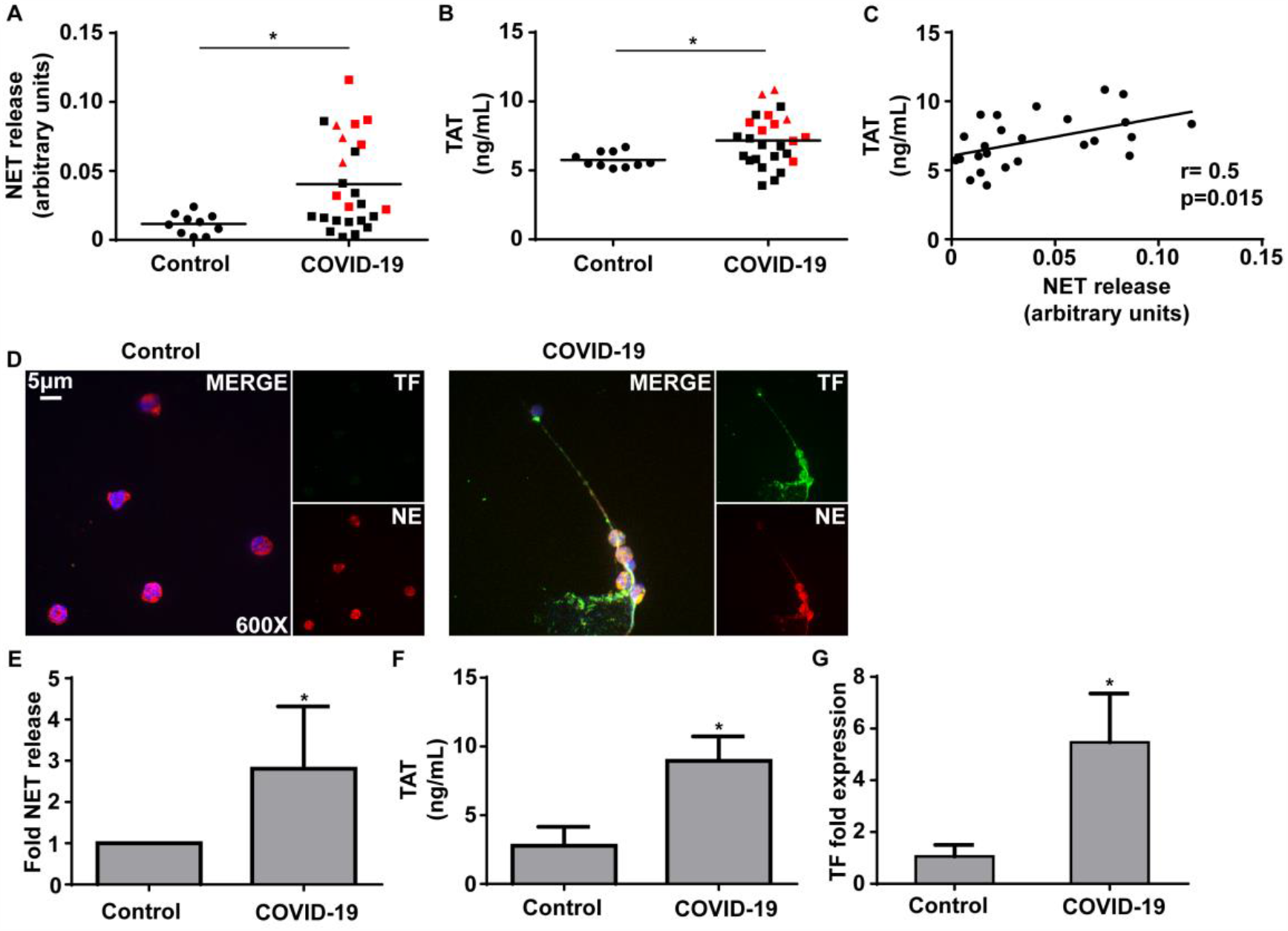
Neutrophil extracellular traps (NETs) in the coagulopathy of COVID-19. **A)** Myeloperoxidase (MPO)-DNA complex levels representing NET release in plasma from healthy individuals (controls, n=10) and COVID-19 patients (n=25) **B)** Thrombin-antithrombin (TAT) complex levels in plasma from controls and COVID-19 patients. In both A and B, red squares denote patients with severe COVID-19, red triangles denote critical COVD-19 patients. **C)** Correlation between MPO-DNA complex levels and TAT complex levels in COVID-19 patients. **D)** Confocal fluorescence microscopy showing tissue factor (TF)/neutrophil elastase (NE) staining in neutrophils isolated from controls and COVID-19 patients. A representative example of four independent experiments is shown. Original magnification: x600, scale bar: 5 μm. Blue: DAPI, green: TF, red: NE. **E)** MPO-DNA complex levels in NET structures isolated from controls and COVID-19 patients. Data are from four independent experiments (mean ± SD). **F)** TAT complex levels in plasma obtained from controls treated with NET structures isolated from controls and COVID-19 patients. Data are from four independent experiments (mean ± SD). **G)**-Fold expression of TF mRNA in neutrophils isolated from controls and COVID-19 patients. Data are from four independent experiments (mean ± SD). **D-G** were performed in the same four patients (identified by * in Supplementary Table 1). All conditions were compared to controls, and statistical significance at p< 0.05 is indicated by *. A, B: Student’s t-test, C: Spearman correlation test, E-G: Friedman’s test.

In an effort to corroborate our *ex vivo* findings, and because platelet-neutrophil interactions are necessary for NET formation in several thomboinflammatory disorders (10, 18), we used PRP derived from COVID-19 patients to perform *in vitro* stimulation of neutrophils isolated from healthy individuals (control neutrophils). We found that these PRP-stimulated control neutrophils had increased levels of TF mRNA (Figure 2A) and efficiently generated TF-bearing NETs (Figure 2B,D,E); the corresponding NET structures showed high TAT activity (Figure 2C), supporting the important role of platelets in NET-mediated COVID-19 immunothrombosis. On the other hand, although COVID-19 serum or plasma led to intracellular TF expression in neutrophils, they were not able to trigger efficient NET formation (Figure S1A-C), further supporting previous studies indicating that the release of TF-expressing NETs is a “double-hit phenomenon” (10, 19). That is, PRP includes both the first hit, which induces TF expression (plasma), and the second hit, which enables NET formation and extracellular exposure of TF via NETs (platelets).

**Figure 2.**
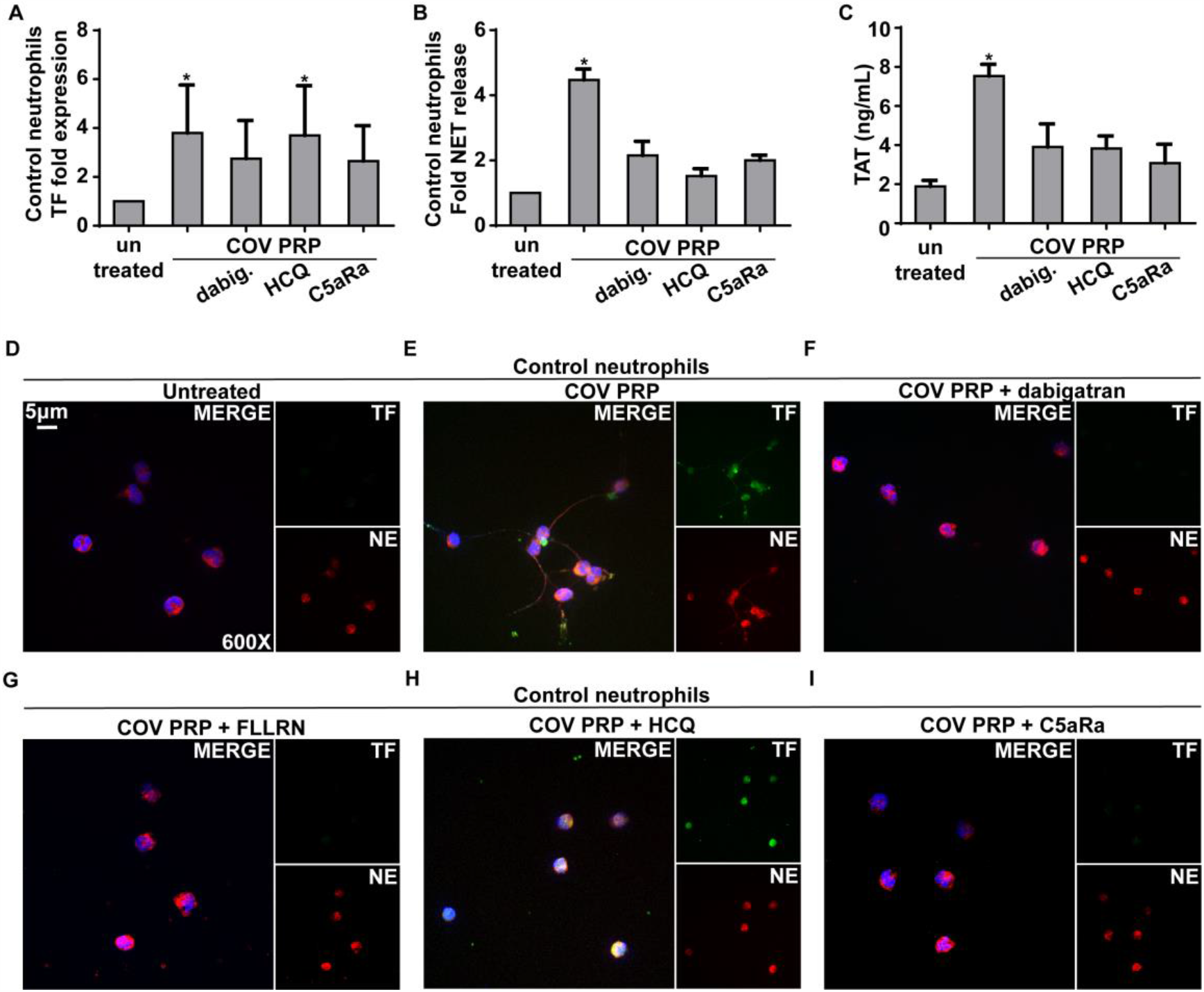
Inhibition studies of platelet-rich plasma (PRP)-neutrophil interactions in COVID-19. **A)**-Fold expression of tissue factor (TF) mRNA in control neutrophils treated with COVID-19-derived PRP (COV PRP) and inhibited with dabigatran (thrombin inhibitor), hydroxychloroquine (HCQ, autophagy inhibitor) or C5aR1 antagonist (C5aRa/PMX-53). Data are from four independent experiments (mean ± SD). **B)** Myeloperoxidase (MPO)-DNA complex levels in NET structures isolated from control neutrophils treated with COV PRP and inhibited with dabigatran, HCQ or C5aRa/PMX-53. Data are from four independent experiments (mean ± SD). **C)** Thrombin-antithrombin (TAT) complex levels in control plasma stimulated with NET structures isolated from control neutrophils treated with COV PRP and inhibited with dabigatran, HCQ or C5aR. Data are from four independent experiments (mean ± SD). **D, E)** Confocal fluorescence microscopy showing tissue factor (TF)/neutrophil elastase (NE) staining in control neutrophils treated with COVID-19-derived PRP and inhibited with **F)** dabigatran, **G)** FLLRN (PAR1 receptor inhibitor), **H)** HCQ or **I)** C5aRa/PMX-53. A representative example of four independent experiments is shown. Original magnification: x600, scale bar: 5 μm. Blue: DAPI, green: TF, red: NE. All conditions were compared to control/untreated; statistical significance at p< 0.05 is indicated by *. A-C: Friedman’s test.

Considering the proposed role of endothelial cells in the thrombotic microangiopathy of COVID-19 (2, 20), we next undertook an *in vitro* investigation of the interplay between HAEC and the platelet/neutrophil/TF axis. Only NETs generated *in vitro* by control neutrophils that had been exposed to PRP from COVID-19 patients were able to induce TF expression in HAEC that led to thrombotic potential, as indicated by TF qPCR (Figure S2A), and TAT assay in cell culture supernatants (Figure S2B). In contrast, although HAEC treated with COVID-19-derived PRP alone expressed endothelial activation markers (Figure S2C), the COVID-19 PRP was not able to stimulate TF activity in the HAEC (Figure S2A,B). Moreover, PMA-induced NETs had no effect on HAEC (Figure S2A-C).

Collectively, these results suggest that COVID-19 PRP-induced NETs may amplify the TF/thrombin axis by activating endothelial cells to express TF.

### Inhibition of NET-driven immunothrombosis

We next expanded our findings by investigating the possible mechanisms governing platelet/NET/TF cross-talk. Given that activated platelets provide a catalytic surface for thrombin generation (21), we examined the role of thrombin in platelet/neutrophil interactions. Thrombin inhibition with dabigatran mitigated TF expression (Figure 2A,F) and activity (Figure 2C), as well as NET release (Figure 2B,F) in COVID-19 PRP-stimulated neutrophils, indicating that thrombin contributes to platelet/neutrophil-mediated thrombogenicity. Similar effects were also obtained by blocking thrombin signaling with the FLLRN peptide against the PAR1 receptor (Figure 2G). These findings are in line with current guidelines recommending the use of low molecular weight heparin (LMWH) in COVID-19 patients (22). Moreover, the intracellular signaling of thrombin through PAR1 implies a potential link with inflammatory pathways (23).

Several reports have demonstrated that autophagy is tightly associated with NET formation (24). Pretreatment of neutrophils with the autophagy inhibitor hydroxychloroquine (HCQ), which is currently being tested in COVID-19 patients (25), abolished NET formation in PRP-stimulated neutrophils (Figure 2B,H), leading to a reduction in NET-bound active TF, as assessed by TAT assay (Figure 2C). This finding suggests that autophagy may be involved in platelet-induced NET formation. It also suggests additional action by HCQ in COVID-19 against immunothrombosis and further supports both earlier and recent data indicating a promising effect for HCQ in complement- and NET-related thromboinflammatory disorders (26–28).

Since C5a is a key mediator of neutrophil TF expression (14–17), we next selectively inhibited its signaling in PRP-stimulated neutrophils from COVID-19 patients. We found that C5a receptor (C5aR1) blockade attenuated TF expression at the mRNA level as well as in immunofluorescent staining for TF (Figure 2A and I, respectively); blocking the C5aRI also inhibited NET release, as indicated by ELISA assay of MPO/DNA complex levels and immunostaining (Figure 2B and I, respectively). Consequently, we also saw diminished TF functionality as assessed by TAT assay (Figure 2C). These findings are consistent with previous studies indicating a dual role for complement activation in both TF expression by neutrophils (14–17) and mechanisms related to NET formation (29, 30). In addition, the COVID-19 patients exhibited considerably increased plasma levels of sC5b-9 (terminal complement complex/TCC) (Figure 3A). Our findings are in agreement with recent evidence demonstrating significantly higher plasma levels of sC5b-9 and C5a in severely affected COVID-19 patients and widespread complement deposition along with microvascular thrombosis in lung and skin tissue samples (9, 31, 32).

**Figure 3.**
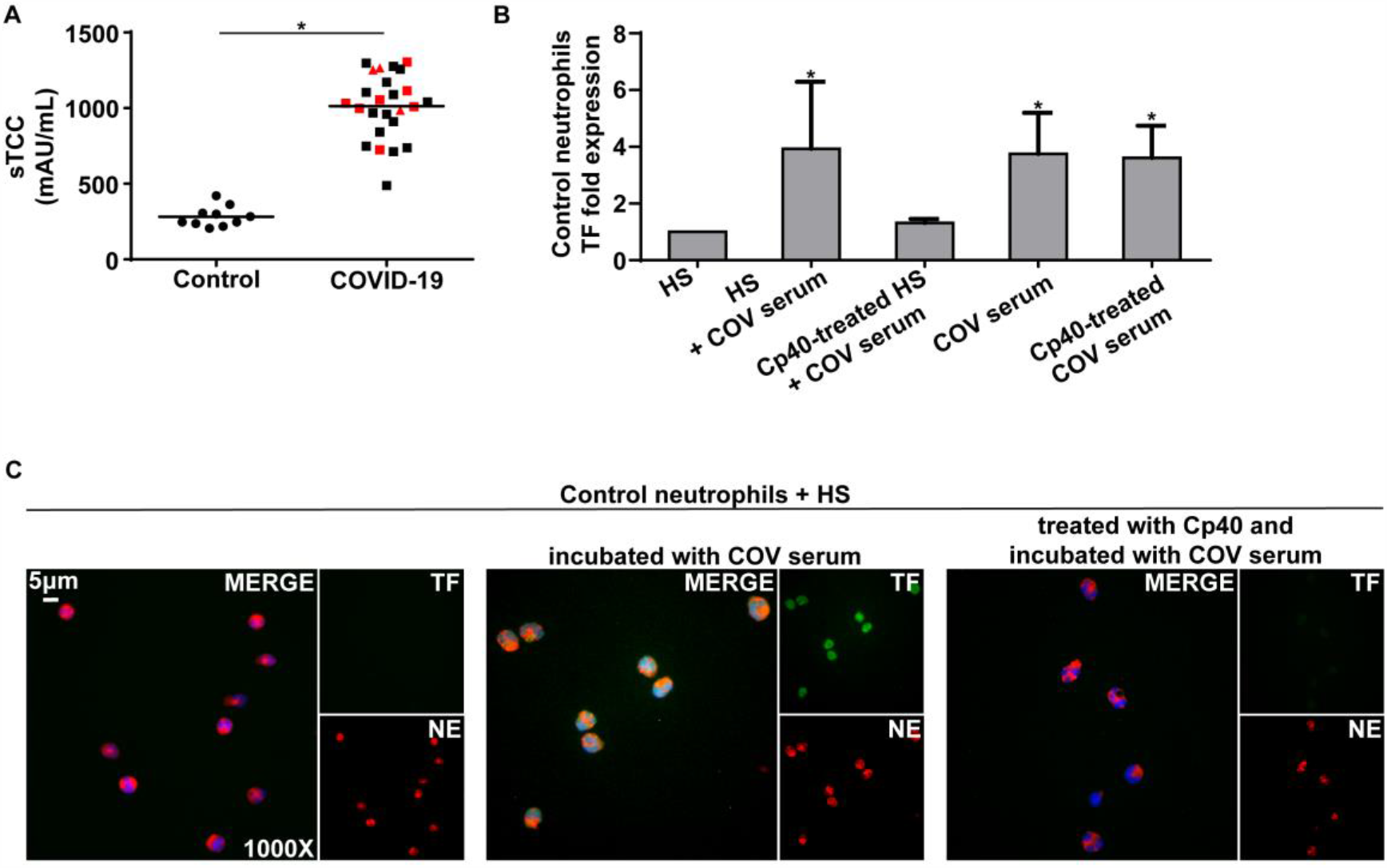
C3 inhibition disrupts neutrophil-driven thromboinflammation in COVID-19. **A)** Soluble terminal complement complex (sTCC) levels in plasma from controls (n=10) and COVID-19 patients (n=25). Red squares: patients with severe COVID-19, red triangles: critical patients. **B)**-Fold expression of TF mRNA in control neutrophils stimulated with: serum from healthy individuals (HS), HS incubated with COVID-19 serum (“COV serum”), HS treated with compstatin analog Cp40 and then COV serum, COV serum alone, or COV serum treated with Cp40. Data are from four independent experiments (mean ± SD). **C)** Fluorescence microscopy showing tissue factor (TF)/neutrophil elastase (NE) staining in control neutrophils stimulated with: HS, HS incubated with COV serum, or HS treated with Cp40 and then COV serum. A representative example of four independent experiments is shown. Original magnification: x1000, scale bar: 5 μm. Blue: DAPI, green: TF, red: NE. All conditions were compared to HS alone (control), and statistical significance at p< 0.05 is indicated by *. A: Student’s t-test, B: Friedman’s test.

### Compstatin inhibits complement activation, disrupting neutrophil-mediated TF expression

Emerging evidence from COVID-19 patients indicates that complement activation predominantly involves the lectin and alternative complement pathways. However, the possible contribution of the classical pathway, due to immune complexes formed by natural autoantibodies or antibodies against SARS-CoV-2, cannot be ruled out (12, 32).

Given the central role of C3 in the complement cascade as a point of convergence of all pathways, it is reasonable to hypothesize that C3 activation is a crucial upstream mechanism driving C5a/C5aR1-mediated responses in COVID-19.

Prompted by our findings described above regarding the high levels of complement activation in COVID-19 patients (increased sC5b-9), we next examined whether COVID-19 patient serum, which represents the disease’s inflammatory environment, is able to induce complement activation upstream of C5, leading to TF expression in neutrophils. In this context, to address the role of C3 activation we developed a serum co-incubation system (for details see Supplemental Methods, section 1.4.1). Serum from healthy individuals (control serum), serving as a source of non-activated complement, was pre-treated or not treated with the compstatin analog Cp40 (which blocks all three pathways of complement activation by targeting the central protein C3) (12, 16), and then co-incubated with COVID-19 serum. Cp40-mediated inhibition of C3 significantly decreased the capacity of our serum co-incubation system to induce TF expression in neutrophils, at both the mRNA (Figure 3B) and protein levels (Figure 3C). These results indicate that C3 inhibition disrupts the source that triggers TF release from neutrophils, broadly preventing complement activation and impairing the thrombogenicity of the TF/thrombin axis in COVID-19.

COVID-19 serum contains a plurality of plausible factors that could trigger *de novo* C3 activation in the control serum; these include immune complexes due to the presence of natural autoantibodies, or cross-reactive IgM recognizing conserved epitopes of ‘common cold’ coronavirus strains, or SARS-CoV-2-induced specific IgG antibodies. Triggers of C3 activation could also include virus-released pathogen-associated molecular patterns (PAMPs) such as the nucleocapsid (N) protein that has been linked to mannan-binding lectin serine protease-2 (MASP-2)-dependent lectin pathway activation (12, 32).

The above finding is further supported by recent, promising clinical data from the compassionate use of the C3-targeted therapeutic AMY-101 (Cp40-based drug candidate, clinically developed by Amyndas Pharmaceuticals) in severely ill COVID-19 patients with ARDS (13). AMY-101 treatment resulted in rapid normalization of inflammatory markers and improvement of respiratory function, indicating a pronounced effect of C3 inhibition on COVID-19 thromboinflammation (13). In this regard, the impact of C3 inhibition on neutrophil-derived TF expression may provide important mechanistic insights into how AMY-101 can broadly suppress the thomboinflammatory response that culminates in pronounced microvascular injury and thrombosis in severely ill COVID-19 patients. A phase II clinical trial has been launched to assess the impact of AMY-101 in patients with severe COVID-19 (NCT04395456).

Many pieces of the immunothrombosis puzzle have been identified as components of the complement/neutrophil/TF axis in various disorders (14–17). In addition, it has recently been demonstrated that C3aR signaling can mediate platelet activation in coronary artery disease (33). The ability of Cp40 to attenuate neutrophil-driven thromboinflammation in our study could reflect a broader impact of C3 activation on neutrophil-platelet interactions that promote thrombogenic responses. In fact, C3 inhibition by compstatin disrupts CR3-dependent platelet-neutrophil complex formation which could likely enhance thrombotic reactions relevant to COVID-19 pathology (34). Moreover, enhanced C3 expression is part of the unique host transcriptional signature in response to SARS-CoV-2 infection (35) and C3aR upregulation in the lung microvascular endothelium was recently correlated with disease severity (36), and increased thrombosis and aberrant vascular angiogenesis in COVID-19 lung biopsies (2). Collectively, these studies support a key role for C3 activation in driving the platelet/NETs coagulopathy axis. Here, we report that this axis is substantially involved in the pathophysiology of COVID-19 (Figure 4). Many questions remain to be answered, such as how the complement system is activated and regulated, how complement over-activation is involved in the aberrant host immune response leading to the COVID-19 cytokine storm, and which components of the NET scaffold in addition to TF are associated with neutrophil driven-thromboinflammation. We propose that COVID-19 constitutes an emerging clinical model of immunothrombosis that may help to shed light on the dark side of complement-related disorders. Our findings argue for the timely application of therapeutic strategies that can disrupt the vicious cycle of COVID-19 immunothrombosis by targeting complement activation or/and NET formation (10, 12, 13, 24, 25, 31, 32) (Figure 4).

**Figure 4.**
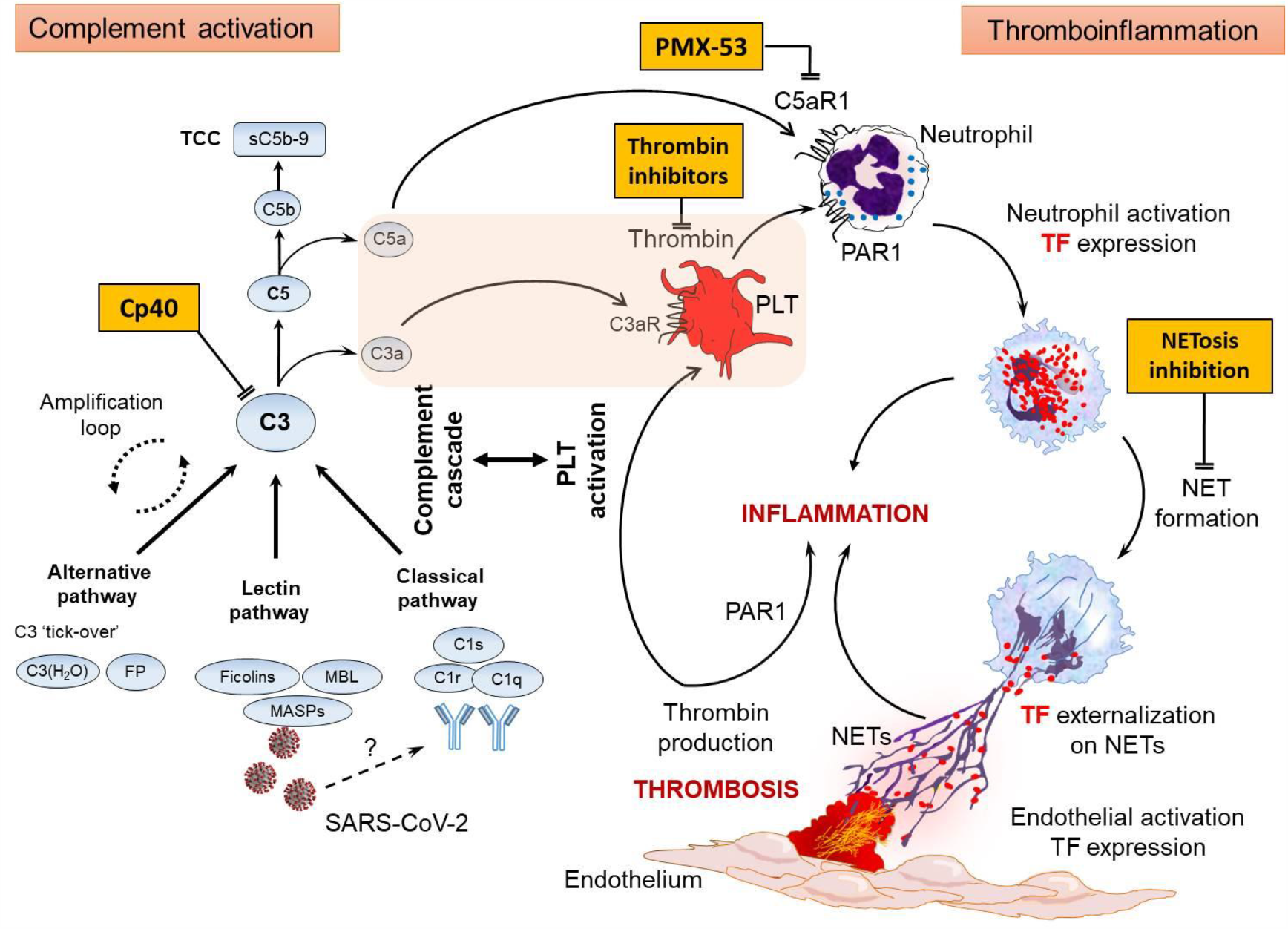
Proposed mechanism of COVID-19 immunothrombosis. During COVID-19, SARS-CoV-2 virus triggers complement cascade by interacting with mannan-binding lectin (MBL) serine proteases (MASPs) or possibly through the induction of (auto)antibodies or/and immunocomplexes. Activation of C3, as a point of convergence of all complement pathways, leads to C3a, C5a and sC5b-9 (terminal complement complex/TCC) generation. Subsequently, C3a activates platelets (PLT), while C5a and PLT-derived thrombin induce both neutrophil tissue factor (TF) expression and neutrophil extracellular traps (NETs) carrying active TF. These thrombogenic NETs may induce endothelial cells activation towards TF expression increasing their procoagulant activity. This may further amplifies (e.g. via PAR1), both inflammation and PLT activation constructing thus a complement/NETs-driven vicious cycle of immunothrombosis. Complement, thrombin and NETosis represent promising candidate therapeutic targets. The pink box includes components of COVID-19 thromboinflammatory environment. Cp40, compstatin analog; FP, properdin; PAR1, protease-activated receptor 1; PMX-53, specific C5a receptor antagonist; SARS-CoV-2, severe acute respiratory syndrome coronavirus 2.

## Methods

The subjects were 25 patients hospitalized at University Hospital of Alexandroupolis or AHEPA University Hospital of Thessaloniki, with moderate (n=15), severe (n=7) or critical (n=3) COVID-19 disease (12 males, 13 females; mean age, 62.1±13.8 years; Supplemental Table S1); 10 healthy age- and sex-matched individuals served as controls. All patients were analyzed 8 to 15 days after disease onset. At the time of sampling, none of the patients were undergoing chemotherapy, radiotherapy, or an immunomodulatory treatment such as tocilizumab, anakinra or corticosteroids. Neutrophils, sera, citrated plasma and platelet-rich plasma (PRP) were isolated from COVID-19 patients and controls. Among the 25 patients of the study, samples from four patients (two males, two females) with severe COVID-19 disease were used for *ex vivo* confocal immunofluorescence microscopy experiments and *in vitro* stimulation studies (indicated by asterisk in Supplemental Table S1). For *in vitro* stimulation, appropriate cultures of human aortic endothelial cell (HAEC) were also used. The study protocol is in accordance with the Declaration of Helsinki and was approved by the Local Ethics Review Board of the hospitals that participated in the study. Written informed consent was received from participants or their relatives prior to inclusion in the study. Detailed information for all methods is provided in the Supplemental Methods.

## Data Availability

All raw data for the generation of graphs will be available if required.

## Author contributions

PS wrote the manuscript, conceived and designed experiments, acquired and analyzed clinical data; AM, AC, designed and conducted experiments, analyzed data and contributed to writing; DCM contributed to data analysis and writing; MN, VT, CT, TC, conducted in vitro experiments; SM, PR, ES, MT, CP, IM, GG provided clinical samples and analyzed data; JDL, KR contributed to writing, conceived, designed and co-supervised the study. All authors have read and approved the final manuscript.

## Acknowledgments

Immunofluorescence experiments were performed at the Cell Imaging and Biomolecular Interactions facility of Democritus University of Thrace, Alexandroupolis, Greece.

We thank Dr. Vasileios Papadopoulos for his advice in statistical analysis and Dr. D. McClellan for editorial assistance.

This study was supported by GSRT, EYDE-ETAK Research & Innovation Programme CYTONET, Grant no: ?1?DK-00617.

AM is co-financed by Greece and the European Union (European Social Fund-ESF) through the Operational Programme «Human Resources Development, Education and Lifelong Learning» in the context of the project “Strengthening Human Resources Research Potential via Doctorate Research” (MIS-5000432), implemented by the State Scholarships Foundation (IKY).

